# Altered resting-state functional connectivity in hiPSC-derived neuronal networks from schizophrenia patients

**DOI:** 10.1101/2021.08.26.21262277

**Authors:** Sofía Puvogel, Kris Blanchard, Bárbara S. Casas, Robyn Miller, Delia Garrido, Magdalena Sanhueza, Verónica Palma

## Abstract

Schizophrenia (SZ) is a complex mental disease thought to arise from abnormal neurodevelopment, characterized by an altered reality perception and widely associated with brain connectivity anomalies. Previous work has shown disrupted resting-state brain functional connectivity (FC) in SZ patients. We used Human Induced Pluripotent Stem Cells (hiPSC)-derived neuronal cultures to study SZ’s neural communicational dynamics during early development. We conducted gene and protein expression profiling, calcium imaging and mathematical modeling to evaluate FC. Along the neurodifferentiation process, SZ networks displayed altered expression of genes related to synaptic function, cell migration and cytoskeleton organization, suggesting alterations in excitatory/inhibitory balance. Resting-state FC in neuronal networks derived from healthy controls (HC) and SZ patients emerged as a dynamic phenomenon exhibiting “hub-states”, which are connectivity configurations reoccurring in time. Compared to HC, SZ networks were less thorough in exploring different FC configurations, changed configurations less often, presented a reduced repertoire of hub-states and spent longer uninterrupted time intervals in this less diverse universe of hubs. Our observations at a single cell resolution may reflect intrinsic dynamical principles ruling brain activity at rest and highlight the relevance of identifying multiscale connectivity properties between functional brain units. We propose that FC alterations in SZ patients are a consequence of an abnormal early development of synaptic communication dynamics, compromising network’s ability for rapid and efficient reorganization of neuronal activity patterns. Remarkably, these findings mirror resting-state brain FC in SZ patients, laying the groundwork for future studies among such different spatiotemporal domains, as are brains and neurons, in both health and disease.

## INTRODUCTION

Schizophrenia (SZ) is a severe and chronic mental disorder that affects over 20 million people worldwide [1]. It is a polygenic disease with multiple etiologies [2], that originates during nervous system development, despite the fact that it is diagnosed during adolescence [3]. Although Bleuler first described it back in the 19th century, to date, the mechanisms triggering and predicting the evolution of the disease remain largely unknown. Different studies correlate SZ symptomatology with an altered communication between brain regions. In terms of structural connectivity, SZ brains show significant reductions in the white and grey matter [4]. Moreover, there is a wealth of evidence showing aberrant functional connectivity (FC) in SZ. FC, defined by Joana Cabral as “the temporal dependence of neuronal activity patterns of anatomically separated brain regions” [5], can be measured across brain with functional magnetic resonance imaging (fMRI) [6]. fMRI results have shown an abnormal brain FC in SZ patients in both resting state [7-10] and during task performances [8, 9, 11]. Many of these studies have assumed that FC is static during the timescale of an fMRI scan (∼30 min) and only recently it became clear that resting-state FC is instead characterized by visiting different time-varying connectivity patterns [5, 12]. This flow through different configurations is not arbitrary since discrete connectivity states consistently recur throughout subject scans [13]. These findings have promoted greater interest for more detailed dynamical analyses of time-varying FC. Evaluation of resting-state FC has shown pronounced reductions in the number and diversity of connectivity states in SZ patients compared to healthy individuals, reflecting alterations in brain communicational dynamics [14]. To date, FC datasets have been obtained via fMRI scanning in adults diagnosed with SZ. fMRI has a limited ability to measure brain activity with high temporal and spatial resolution, since it relies on hemodynamic changes. This technique allows assessing correlations between the signals from different parcellated brains nodes (voxels). A typical fMRI voxel size is around 3×3×3 mm^3^, reflecting the average activity of hundreds to thousands of neurons [15]. Such volume covers the entire cortical thickness, containing several cell types with different morphological and functional properties. Hence, there is still a gap in the understanding of the neurobiological processes underlying FC anomalies in SZ.

As synapses are the elementary functional units allowing information flow between neurons, studying neuronal communication during development could significantly increase our understanding of SZ connectivity disfunction. Human induced pluripotent stem cells (hiPSC) models rescue the whole genetic component of patients with complex polygenic brain disorders [16] such as SZ. These cells can faithfully recapitulate neurogenesis and the evolution of spontaneous network activity during brain development [17-22]. Therefore, these cultures represent a novel strategy to study developmental psychiatric diseases. While it is not possible to reproduce the complexity of specific brain regions and their connectivity, this *in vitro* model may allow exploring general FC dynamical properties of neuronal networks in resting conditions. This approach could shed light on intrinsic connectivity alterations in SZ at a single-cell resolution.

With this aim, we focused on quantifying the dynamics of FC in long-term hiPSC-derived neuronal networks of SZ patients and healthy control subjects (HC). Using Ca^2+^ imaging, we studied network spontaneous activity and evaluated the temporal correlation among Ca^2+^ transients’ patterns in these large neuronal populations. We adapted an analytic method previously designed to evaluate whole-brain high-level FC dynamics [14], to work at a cellular level. We quantified the competence of these networks for rapid connectivity reshaping and their ability to explore different functional configurations, among other dynamic properties.

We found alterations in FC dynamics of SZ networks emerging from hiPSC models resembling early fetal stages. Remarkably, our approach allowed recapitulating *in vitro*, at the cellular level, the functional dynamics observed by whole-brain fMRI scanning. These observations might have global implications in our understanding of communicational properties of SZ brains. Furthermore, they may open new routes for exploring the application of fundamental dynamical principles to more complex levels of organization in both normal and pathological situations.

## METHODS AND MATERIALS

### Differentiation of NSC to mature neuronal cultures

hiPSC-derived NSC from 3 healthy subjects and 4 SZ patients were kindly donated by Dr. Stevens Rehen at D’Or Insitute and Feredal Univeristy of Rio de Janeiro. Differentiation of hiPSC to NSC was performed as previously published [23]. The establishment of hiPSC, and derivation of NSC lines were carried out in accordance with international standards and with the approval of the Research Ethics Council “Comité de Ética em Pesquisa – Hospital Copa D’Or, Rua Figueiredo Magalhães 875, Rio de Janeiro, Brasil.” (Certificado de Apresentação de Apreciação Ética (CAAE): 32385314.9.0000.5249). Detailed information on patient samples and reprogramming protocol can be found in Table S1. NSC differentiation to mature neuronal networks was conducted adapting a protocol of Shi et al. [24]. NSCs were thawed and seeded in Geltrex-coated plastic 60 mm plates and maintained in Neural Expansion Media (NEM; DMEM/F12 and Neurobasal medium (1:1) with Neural Induction Supplement; Thermo Fisher Scientific, Carlsbad, CA, USA) until they reached 80-100% confluence. Next, NSC were detached from the plate with accutase (7 min/37ºC), centrifugated at 300*g* for 4 min, and resuspended in NEM plus 10 µM Rock Inhibitor (Y-27632; Merck, Darmstadt, Germany). 1*10^6 cells were plated in 35 mm petri dishes coated by poly-l-ornithine/laminin (10 μg/ml and 2.5 μg/ml respectively). After 48 h, the medium was replaced by Neural Differentiation Media (NDM; DMEM/F12 and Neurobasal medium (1:1) supplemented with 1×N2, 1×B27, 2-Mercaptoethanol 100uM). NDM was changed every two days. 14-16 days after plating, cells were detached from the plate with accutase and passed in a 1:3 ratio into 35 mm poly-l-ornithine/laminin-coated plates. Medium changes were done every 2 days. Around day 30, cells were passed into their final coated 35mm plate, in a 1:4 ratio (around 500.000 cells per 35 mm petri dish). Cells were maintained in NDM, with medium changes every other day, for 60 more days and were supplemented with 10 µM Rock inhibitor for 48 h, after every passage.

### Immunofluorescence

At 60 days of differentiation, cells were fixed with paraformaldehyde 4%, permeated with Triton-X (Sigma, St. Louis, MO, USA) 0.2%, blocked with BSA 5% in PBST (tween-20 0.1% in PBS) solution, and incubated overnight at 4ºC with anti-tau1 (mouse; #MAB3420 Milipore), anti-synaptophysin (mouse; #101011 Synaptic Systems), anti-MAP2 (rabbit; #AB5622 Millipore), anti-homer-1 (rabbit; #160002 Synaptic Systems). Subsequently, cells were incubated with secondary antibodies (goat anti-mouse alexa 488, goat anti-rabbit alexa 555, Invitrogen). Nuclei were stained with 4′-6-diamino-2-phenylindole (DAPI) 1µg/mL for 5 min. Images were acquired with a Zeiss LSM 710 bi-photonic confocal microscope.

### Electrophysiology

Whole-cell voltage-clamp recordings were performed at 60 days of differentiation in HC and SZ-derived cultures. Neurons were selected following morphological criteria. Na^+^ inward and K^+^ outward currents were evoked by voltage steps ranging from -100 to +40 mV in 10 mV increments (V_hold_ = -75 mV). Data was acquired at 50 kHz and low-pass filtered at 2.9 kHz, by an EPC-10 amplifier (HEKA Elektronik GmbH, Reutlingen, Germany). Patch electrodes (∼4.5 MΩ) were pulled from borosilicate glass and the intracellular solution contained (in mM): 135 K-gluconate, 2 MgCl_2_, 2 Na_2_ATP, 0.3 NaGTP, 10 HEPES, 7 NaCl (pH 7.4). The recording chamber was continuously perfused at 1–2 ml/min with ACSF solution containing (in mM): 115 NaCl, 2.5 KCl, 1.3 NaH_2_PO4, 26 NaHCO_3_, 25 glucose, 5 Na-Pyruvate, 2 CaCl_2_ and 1 MgCl_2_ (300 mOsm/kg), gassed with 5% CO_2_/95% O_2_ (pH 7.4). Recordings were performed at room temperature (20–25 °C). Series resistance (10–40 MΩ) was not compensated and recordings were discarded for variations higher than 20% along the experiment.

### qPCR

RNA was obtained from early differentiated cells (30 days) or late differentiated cells (range of 70-91 days, herein defined as 90 days) and stored in Trizol (Thermo Fisher Scientific, Carlsbad, CA, USA) at - 80ºC until further use. For each line donor, three SZ (#1,3-4) and three HC (#1-3) samples from independent differentiation procedures were pooled at each time frame to extract total RNA. cDNA was synthesized from 1 μg RNA, using a M-MLV reverse-transcription kit (Promega, Madison, WI, USA). Primers were specifically designed to measure expression of the genes listed in Figure S1A. Relative gene expression was assessed by qPCR (Agilent Technologies Thermocycler, Santa Clara, CA, USA). mRNA levels were calculated as the fold-change expression via 2^-ΔΔCt^ and gene expression was normalized to three different housekeeping genes *(B2M, 18S, GAPDH)*. Fold*-*changes were assessed relative to control sample or differentiation time.

### Conditioned medium (CM) collection and neuro-proteomic profile

The presence of 30 different neuronal growth factors in serum-free 48 h collected CM (four SZ (#1-4) and three HC (#1-3)) was evaluated from 75 days cell cultures using the Human Neuro Antibody Array II (#ab211063, Abcam, Cambridge, UK). Spots were detected by chemo-luminescence and intensity quantified by densitometry (ImageJ, NIH, USA). The levels of each factor were measured in duplicate and normalized to internal controls provided by the assay.

### Statistical Analysis for mRNA expression and neuro-proteomic profile comparisons

mRNA fold-change normality was assessed with D’Agostino-Pearson and significance of mRNA was determined by a Nested t-test. A linear mixed model with a random intercept for cell-line id was performed to test between- and within-group differences in secreted proteins measured with the neuro-proteome array. Significance of the regression coefficients associated to the group, we used a Z test and Bonferroni correction for multiple comparisons. Statistical significance was set at p < 0.05.

### Calcium imaging

Ca^2+^ transients were recorded at a single-cell resolution at up to 90 days (range of 80-90 days) of differentiation. Three SZ (#2, 3 and 4) and two HC (#1 and 2) cell lines were analyzed (2-3 plates per cell line; 3-27 networks per plate). We used the cell-permeant Ca^2+^ indicator Oregon Green™ 488 BAPTA-1 (OGB-1 AM), (Thermo Fisher Scientific, Carlsbad, CA, USA). The loading solution consisted of OGB1 3.2 µM, Cremophor EL (Merck, Darmstadt, Germany) 0.01% v/v and Pluronic F-127 (Merck, Darmstadt, Germany) 0.4% in NDM. Cells were incubated at 37°C and 7% CO_2_ in the dark for 1 h. After washing twice with NDM, cells were maintained for 30 min before imaging in ACSF (∼ 4.7 min at 10 Hz). In some experiments, TTX (0.2 µM) was added to the bath solution to confirm the AP-dependence of Ca^2+^ transients.

### Imaging analysis

#### Data pre-processing

Bleach correction was conducted for all recordings with the ImageJ Bleach Correction Macro Package [25]. Contrast and brightness were adjusted manually by using imageJ tools. Motion correction and active neurons identification were performed with the CaImAn Constrained Nonnegative Matrix Factorization algorithm [26]. Ca^2+^ transients (ΔF) were obtained by subtracting baseline florescence (quantile 8) and a ΔF matrix was built (rows represent the different neurons and columns the Ca^2+^ signals at a given temporal frame).

#### Network topology analyzes

The Pearson’s correlation index between the signals of every pair of neurons (ΔF matrix rows) was calculated. A FC matrix (neuron id, neuron id) was created from the correlation results for each network. Two neurons were considered as connected if the absolute value of the correlation index was larger than 0.4.

#### Frequency distribution of neuron connectivity degree

To evaluate if the number of neuronal functional connections (connectivity degree) followed a Poisson or Binomial distribution, nonlinear least-squares analysis was performed using SciPy library. The logarithm (log10) of the connectivity degree versus the logarithm of its frequency was plotted and a linear regression was deployed to derive the slope estimate of the power-law fitting. To evaluate between- and within-group differences in scaling exponents, we used the linear mixed model described below, with a random intercept for cell line identity (id) and by adjusting for the number of neurons and the number of total possible connections in each network.

#### Functional connectivity dynamics

A sliding-time-window correlation method was applied to ΔF matrix [5]. The Person’s correlation index was calculated for the signals of every pair of neurons within a sliding window of 10.5 s, comprising 70 consecutive frames, from frame *t* to frame *t+70*. The window was next shifted in one step, to frame *t+1*, repeating the procedure until all frames were covered. Hence, one FC matrix was obtained per step (wFC(t)). As matrices are symmetrical, we reshaped them into a one-dimensional vector containing just the values below the diagonal. FCD matrices (Figure 3A) were then obtained by computing the Pearson’s correlation coefficient between every pair of wFC(t) vectors.

#### Functional connectivity meta-states

For each network, we conducted independent component analysis (ICA) among the whole set of wFC(t) vectors, using the FastICA algorithm [27]. The number of independent components was set to four, thereby allowing algorithmic convergence in most recordings, and only those recordings in which the algorithm converged were considered in the analysis. Each wFC(t) vector was regressed on the four independent components or “correlation patterns” (CP). This way, vector dimensionality was reduced to four and the original vector was now described by the four corresponding regression coefficients or “weights”. To group similar wFC(t), we discretized the weights into quartiles, which resulted in a finite number of discrete connectivity states (meta-states). As weights could be positive or negative, we treated them independently during the discretization step. We replaced CP weights by a value in ±{1,2,3,4}, according to its signed quartile. Thus, a meta-state was described by the combination of the four discretized weights. After obtaining the set of FC meta-states per network, we calculated the following dynamical variables describing network connectivity:

- Number of visited meta-states: total number of meta-states realized by each network.
- Number of change-points: number of switches from one meta-state to another.
- Mean time in a meta-state: mean number of consecutive frames that the network remains in a meta-state, translated to units of time.
- Maximum distance between successive meta-states: maximum Manhattan distance between two consecutive meta-states.
- Traveled distance: summation of the distances between successive meta-states along the whole recording time.
- Dynamic range: Manhattan distance (L1) between the most distant meta-states.
- Number of hub meta-sates: total number of meta-states visited at least twice during the recording time.
- Mean time in a hub: mean number of consecutive frames that the network remained in the same hub meta-state, converted to units of time.
- Number of visits to hub states: number of times that the network visited any hub meta-state.

The number of frames was translated to time units by multiplying by the camera delay (∼ 0.1 ms). Imaging analysis was performed by a customized code written in Python. Repeated measurements of the different FC-related variables were obtained for each cell line and the “SZ effect” was evaluated by a mixed linear regression model, with a random intercept for each cell line identity (id) (for more details about models, see Table S2). As in most cases, FC variables correlated to the number of active neurons (see Results), we used deviance statistics to evaluate the necessity to adjust for the number of neurons and number of possible connections. In such cases, the variables were previously centered. We used the Statsmodels package from python to fit the following model:

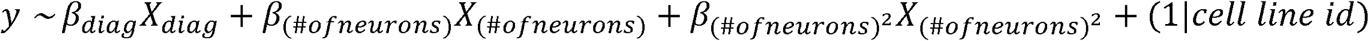

*β*_*i*_ are the coefficients associated to the *X*_*i*_ predictors. *Diagnosis (diag)* is a binary variable, coded as “1” for SZ neurons and as “0” for HC ones. The “SZ effect” corresponds to the value of *β*_*diag*_ when its Z-score associated p-value is less than 0.05. Hence, β_diag_ > 0 indicates a positive correlation of the variable *y* with the SZ condition, and the opposite, for *β*_*diag*_ < 0.

## RESULTS

### Molecular profiling of long-term hiPSC-derived neuronal cultures from SZ and HC subjects

Given the relative scarcity of studies on functional maturation in long-term hiPSC-derived neuronal cultures over time, we first aimed to validate and characterize the differentiation process. We used hiPSC-derived Neural Stem Cell (NSC) cultures from HC and SZ patients, displaying clear staining for NESTIN and PAX6, the two most well-known markers for NSC [23]. We modified the protocol from Shi et al., 2012 [24] to induce long-term neuronal differentiation from four SZ and three HC NSC lines (see Table S1 for details). We omitted the use of antibiotics, as they may affect neuronal excitability [28] and modify the expression of several of genes [29]. HC and SZ patient-derived hiPSC aggregated and formed neuronal rosettes that became larger and more defined along the differentiation process [21]. After 30 days, 3D neuronal aggregates were already easily identified (Figure 1A-D). At 60 days, neurons expressed specific dendritic and axonal markers (MAP2 and TAU, respectively); MAP2 was expressed in perikarya and dendrites (Figure 1E, G), while TAU was mainly found in axons (Figure 1F, H). The pre and postsynaptic proteins synaptophysin (SYP) (Figure 1E, G) and HOMER1 (Figure 1F, H), respectively, were also detected, confirming the presence of synaptic structures. Moreover, both HC and SZ hiPSC-derived neurons displayed voltage-dependent Na^+^ and K^+^ currents (Figure 1I, J).

**Figure 1.**
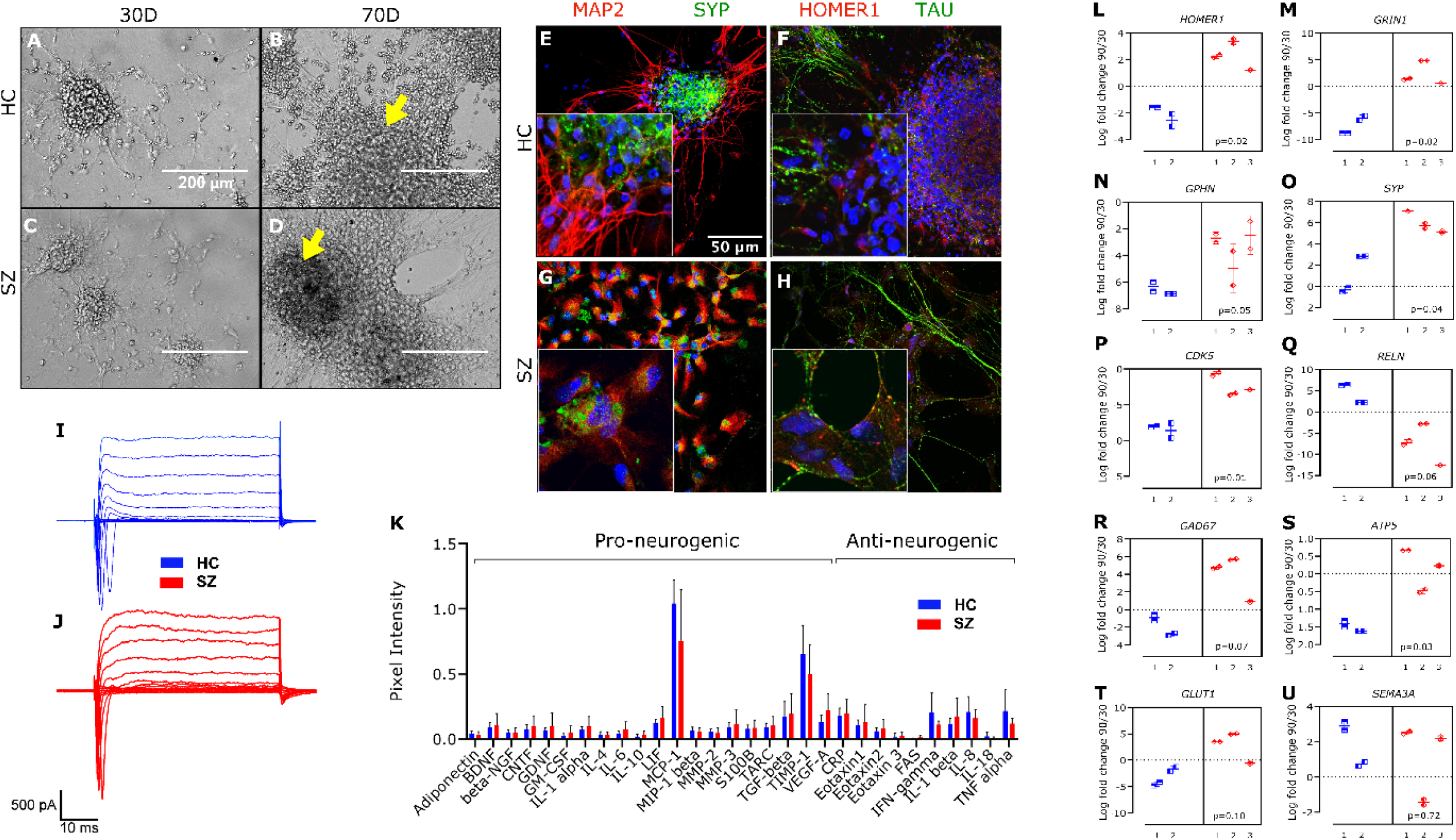
Profiling of hiPSC derived neurons obtained from HC and SZ patients. (A-D) Representative images for HC (A, C) and SZ (B, D) derived neuronal cultures, captured with phase-contrast microscopy at 30 (A, C) and 90 (B, D) days of differentiation. Arrows indicate 3D neuronal aggregates where spontaneous activity was recorded. Calibration bar: 200µm. (E-H) Representative immune staining for the dendritic and axonal markers MAP2 (E, G) and TAU (F, H), respectively, as well as for the pan-presynaptic and glutamatergic-postsynaptic proteins synaptophysin (SYP) (E, G) and HOMER1 (F, H), respectively. DAPI staining is shown in blue. Calibration bar: 36 µm. (I, J) Representative voltage-clamp electrophysiological recordings at 60 days of differentiation, for HC and SZ-derived neurons. (K) Quantification of neurogenic protein levels (mean ± SD) in the CM of four SZ (#1-4) and three HC (#1-3) cell lines at 75 days of differentiation. (L-U) Fold-change in mRNA expression levels of different genes related to central nervous system development, in the period from 30 to 90 days. Data is presented as the log2 of the 90/30 days ratio for three SZ (#1, 2, and 3) and two HC (#2 and 3) cell lines. *B2M* was used as housekeeping; *p <0.05, nested t-test.

The functional competence of our differentiated cell cultures was further assessed by analyzing their secretome. The expression profile of different cytokines/growth factors related to neuronal differentiation and signaling was evaluated by collecting the serum-free CM) from the SZ and HC cell lines (Figure 1K). Quantification of secreted proteins at 75 days of differentiation revealed a similar profile; the complete set of targeted proteins was identified (although Eotaxin-3 and Interleukin-18 were barely detected; Figure S2). Altogether, these results validate the correct establishment of HC and SZ neuronal cultures.

### Altered expression of genes involved in synaptic function and network establishment during SZ neurodevelopment

We analyzed the expression of a set of genes reported to be altered in SZ. Gene expression was quantified by qPCR at two different time points during the neurodevelopmental process: 30 and 90 days in culture (Figure S1A). First, we evaluated genes encoding selected synaptic proteins, including the glutamatergic NMDA-receptor subunit 1 GRIN1 and the postsynaptic scaffolding protein HOMER1 [30]; as well as the GABAergic postsynaptic anchoring protein GEPHYRIN [31]. We also assessed the gene coding for the pan-presynaptic vesicle-associated protein synaptophysin SYP. SZ and HC synaptic phenotypes do not display significant disparities for these genes when compared at the same culture age (Figure S1B). To explore how glutamatergic and GABAergic synapses evolve over time in both conditions, we quantified gene expression change between 30 and 90 days *in vitro* (90/30 days ratio; Figure 1L-O). For details on statistical tests and p-values, see Methods and Materials, Figure 1 and Table S3). Notably, *HOMER1* evolution was significantly different among both conditions (Figure 1L); while expression decreased in HC lines, it increased in SZ. A difference among groups was also detected for *GRIN1* (Figure 1M), with a significant decrease in HC and a trend to increase in SZ. These results point to a divergent evolution in the number or strength of excitatory synapses during early neurodevelopment. On the other hand, expression of the GABAergic marker *GPHN* was reduced in both conditions along this period (Figure 1N), suggesting a decline in GABAergic communication over time. However, the reduction was significantly lower in SZ. Differences in *SYP* expression (Figure 1O) probably reflect the combined effect on excitatory and inhibitory synapses, further supporting an altered evolution of synaptic connectivity over time in SZ compared to HC networks. Overall, these results are consistent with previous hypotheses proposing an altered excitatory/inhibitory balance in SZ brains [32] and suggest that these abnormalities may originate at early stages of neurodevelopment.

We also evaluated expression changes over time of different genes implicated in neurodevelopment, as *CDK5R1* and *REELIN* (*RELN*). *CDK5R1* codes for p35, the neuronal-specific activator of cyclin-dependent kinase 5 (CDK5) regulatory subunit 1 [33]. CDK5 is involved in orienting neuronal network structure and regulating activity during neurodevelopment, via cytoskeleton remodeling [34]. Expression of p35, and hence CDK5 activity, varies cyclically along brain development [35]. During the assessed period, we found a reduction of *CDK5R1* expression in both conditions (Figure 1P, Table S3). Nonetheless, this decrement was significantly lower in SZ than in HC cultures (Figure 1P). The extracellular-matrix glycoprotein *REELIN* plays a critical early role in neuronal migration [36]. Later, it modulates dendritic and axonal outgrow, and spine maturation, by regulating the cytoskeleton. Interestingly, while *RELN* did not significantly change in HC in the observed period, it decreased in SZ (Figure 1Q, Table S3). Glutamate decarboxylase GAD67 is involved in GABA synthesis, but for neuron properties no directly related to neurotransmission, as synaptogenesis and neuroprotection [37]. In contrast to HC, we detected a significant increase in *GAD67* in SZ along the evaluated period (Figure 1R, Table S3).

Since metabolic anomalies have been associated to SZ [38], we also evaluated the expression of *ATP5*, coding for mitochondrial-membrane ATP synthase. *ATP5* expression decreased in the same time interval in HC networks, but did not change in SZ ones (Figure 1S, Table S3). An abnormal energy metabolism during early neurodevelopment might have multiple consequences on network establishment and function [39].

Finally, we did not detect changes in the glucose transporter 1 gene (*GLUT1*) and the member of the semaphorin family *SEMA3A* along time (Figure 1T-U).

### Calcium imaging in hiPSC-derived neuronal networks

Previous studies have assessed neuronal structural connectivity in hiPSC-derived neurons using trans-neuronal spread of rabies virus [19, 40]. Neuronal cultures derived from SZ patients presented a decrease in transsynaptic tracing [40], which could be due to a decrement in the number of synaptic connections. However, electrophysiological recordings revealed similar spontaneous synaptic activity in SZ and HC cultures [40]. The activity of neurons triggering action potentials can be monitored by sensing changes in intracellular Ca^2+^ concentration [41]. To address neuronal network connectivity from a functional perspective, we recorded the spontaneous activity of hiPSC-derived populations of neurons at a single-cell resolution and quantified the co-variation of activity patterns between each pair of neurons (3-27 networks per plate, 2-3 plates per cell line, 3 SZ and 2 HC cell lines, Table S1).

Cell cultures were loaded with the cell-permeant Ca^2+^ indicator OGB1 (Figure 2A, B). We recorded 4.7 min length videos (10 pictures/second) of different regions of interest (ROI) within the cell plates, while illuminating with blue light (OGB1 peak absorption = 493 nm). Spontaneous activity was found with a higher probability in neuronal aggregates protruding from the base of the plate (Figure 1B, D; black arrow), as previously described [21]. Since each local network had a unique structure, varying in size and shape even within each culture plate, we considered these aggregates as the ROIs to study. All measurements and analyses were conducted in these 3D neuronal aggregates. We identified the active neurons within the ROI using the open-source tool CaImAn software, considering a signal-to-noise ratio above 2.5 [26]. The set of active neurons within an aggregate was defined as a neuronal network. Ca^2+^ transients (ΔF) were quantified from large populations of cells (Figure 2C, D). Action potential-evoked Ca^2+^ transients depended on the activation of voltage-gated Na^+^ channels as their frequency was strongly decreased after adding tetrodotoxin (TTX; 0.2 µM) to the bath during recordings (reduction of about 43% in the number of active neurons, from 240 to 137; Figure S3).

**Figure 2.**
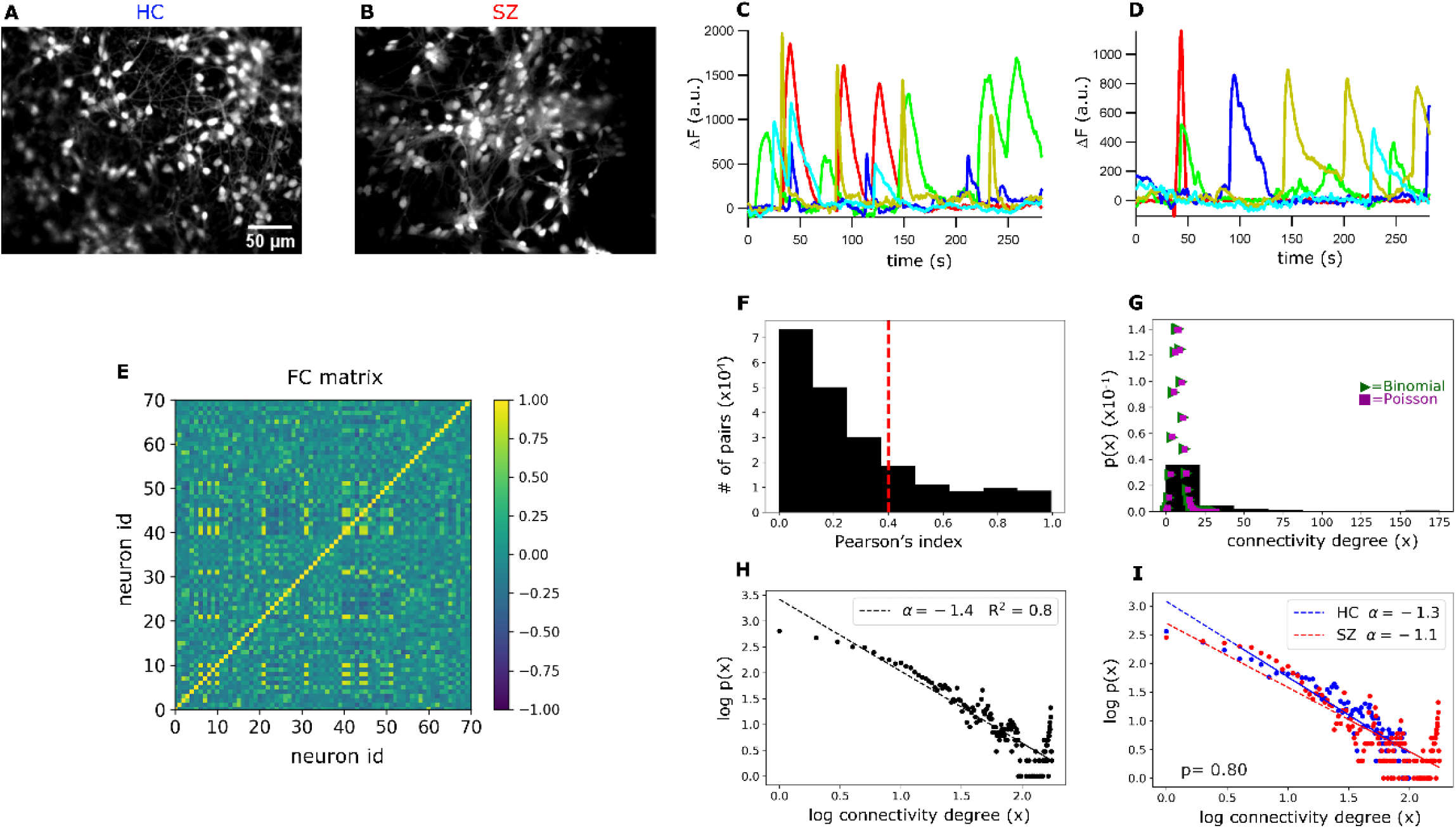
Functional connectivity (FC) and topology analysis in HC and SZ networks. (A, B) Representative ROI images of HC and SZ neuronal cultures loaded with the Ca^2+^ indicator OGB-1. (C, D) Ca^2+^ fluctuations (change in fluorescence intensity relative to baseline (80% of the signal)) in 5 randomly selected neurons from an HC and a SZ network. (E) Static FC matrix displaying the average Pearson’s correlation index (color bar) between Ca^2+^ signals from each pair of neurons in an HC network. (F) Frequency plot of Pearson’s correlation coefficients among neuron pairs from all networks. The dotted red line represents the threshold beyond which we considered a pair of neurons as functionally connected. (G) Probability density of finding a neuron with “x” number of functional connections (connectivity degree). The connectivity distribution does not fit to a Poisson (λ = 8) nor to a Binomial (n = 1000, p = 0.008) distribution. (H) Plot of the connectivity-degree logarithm (log10) versus the probability logarithm. Data fits a scale-free or long-tailed distribution, with a scaling exponent of -1.4. (I) Scale-free fitting of neurons connectivity degree in SZ and HC networks. Connectivity scaling exponents were fitted with a mixed effect regression model detailed in Methods. The scaling exponents do not significantly differ between the two groups.

The number of active neurons within each network varied between 15 to 250 and regression analysis showed no difference in the mean number of active neurons per network between SZ and HC conditions (Figure S4A, mixed regression model detailed in Methods). However, because in most cases the measured FC-related variables (see below) correlated to the number of active neurons (Figure S4B), we considered this parameter as a covariable for almost all the multiple regression analyzes.

### Network topology analysis in hiPSC-derived neuronal cultures, assessed by FC measurements

Since the underlying network topology frames neuronal interactions and signaling, assessing the FC properties of these systems is an indirect way of revealing and understanding the network’s structure and a direct way to interrogate the emerging network functioning.

To quantify network FC, we computed the Pearson’s correlation index for the Ca^2+^ signals of every pair of neurons in each network. As a first approach, we considered the entire recording time (4.7 min) and generated a FC matrix (or correlation matrix) displaying correlation indexes among neuron pairs (Figure 2E). A pair was considered as functionally connected when the correlation index was above 0.4 (defined here as positive connections) or below -0.4 (negative connections). For an absolute correlation value of 0.4 as a cut-off point, the functionally connected neurons reached 25% of the total possible connections, considering all networks (Figure 2F). The number of functional connections per neuron was obtained from the FC matrix. If the total number of connections per neuron (including positive and negative connections) were randomly determined (Figure 2G), the probability for a neuron to have “x” connections (connectivity degree) should follow a binomial distribution and for large enough populations, a Poisson distribution. In contrast, the probability distribution of the number of connections per neuron was right-skewed, resembling a ‘scale-free’ distribution (Figure 2H; also known as ‘long-tailed’ or ‘power-law’ distribution). This means that a large number of neurons have few active connections while a small number are hyper-connected (Figure 2G, H, I). This architecture suggests a complex modular organization that may optimize information processing and network robustness to damage [42] [43].

The scale-free fitting of neurons connectivity degree follows a power-law with a scaling exponent of -1.4 (Figure 2H). Interestingly, using trans-neuronal spread of rabies virus, a previous analysis of network topology in hiPSC-derived neurons demonstrated that the number of structural connections of these neurons also follows a scale-free distribution, with power -2 [19]. The latter supports the possibility that, through measures of FC, it is possible to explore the physiological properties of these neuronal networks, establishing a link between anatomical and functional connectivity.

To evaluate differences in functional topology between SZ and HC, we fitted a mixed linear model for the networks power-law scaling exponents, including a varying random intercept for cell line identity (id) and adjusting for the number of neurons and total possible number of connections in each network (see Methods). The functional topology of SZ networks does not deviate significantly from the HC networks behavior (Figure 2I).

### FC in hiPSC-derived neuronal networks is dynamic and goes through reoccurring configurations

The FC analysis of resting networks described so far represents a measure of the average connectivity among pairs of neurons along the whole recording time (4.7 min). However, this approach does not allow to explore dynamic fluctuations of network connectivity that could occur within shorter periods of time.

To assess the temporal dimension of the FC in the hiPSC-derived networks, we applied a sliding-time-window correlation method [5] (Figure 3B.I; window width: 10.5 s or 70 frames) and computed the correlation matrix for every pair of time windows (wFC(t)); Figure 3B.II; custom code written in Python), obtaining a time-versus-time functional connectivity dynamics matrix (FCD) (Figure 3A). Matrix diagonal results from comparing the same time points, thus displaying full correlation. Points located adjacent to the diagonal (Figure 3A, white dotted squares), are also expected to present high correlation since they correspond to nearby, partially overlapping time windows. However, the squared blocks far away from the diagonal (Figure 3A, red dotted squares) suggest the reappearance of a previously visited FC configurations in a distant time point. This reveals that, during the registered time, network resting-state FC went through numerous and reoccurring configurations, rather than evolving in an arbitrary way. Remarkably, this observation resembles previously described FC dynamics results in human brains at resting conditions [13].

**Figure 3.**
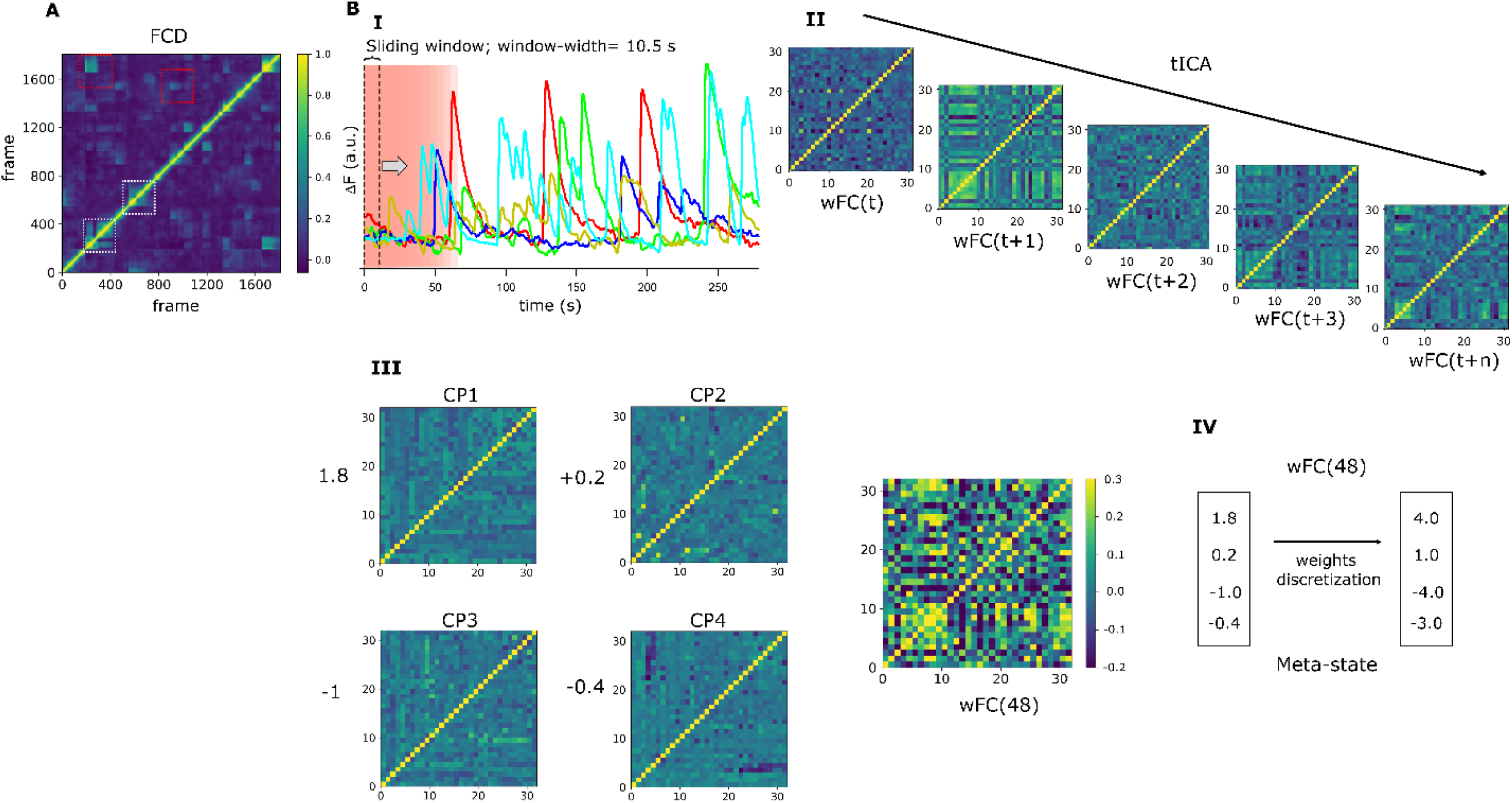
Dynamics of the resting-state functional connectivity (FC). (A) A representative Functional Connectivity Dynamics matrix (FCD) was obtained from an HC network. This time-versus-time FCD(i,j) matrix is obtained by computing the Pearson’s coefficient of every FC(i) centered at a time “i” with every FC(j) centered at time “j”. Periods of lasting FC patterns appear as square blocks around the FCD diagonal (white dotted squares) and reoccurring patterns by square blocks distant from the diagonal (red dotted squares). (B) Scheme illustrating how FC meta-states (MSs) were obtained from an HC network. (B.I) ΔF is computed for every neuron in the network by subtracting the neuron’s baseline signal from the signal recorded at each time point. This plot is a graphical representation showing ΔF for 5 randomly selected neurons within the network, made up by 32 active neurons. A correlation matrix of neuron’s Ca^2+^signal is calculated within a given time-window (t) of shape (n, window width; n: number of neurons, window width = 70 frames = 10.5 sec). Then, the time window is shifted to time t+1 and the correlation matrix is recalculated. The last is successively repeated over the whole recording time. With this procedure, a windowed FC matrix (wFC(t)), with shape (n,n) is obtained for every time step. (B.II) Next, an independent component analysis (tICA) is performed along the temporal axis of the whole set of wFC(t), for every recorded network, and four independent connectivity patterns (CP) are obtained (B.III). Every wFC(t) can be described as a linear combination of these four CP and (B.III) shows the wFC(48) as an example. The coefficients multiplying each CP are obtained by linearly regressing the wFC(48) on the CPs. (B.IV) As we want to have a finite number of states (meta-states), the CP’s coefficients for each network were discretized and assigned to its respective quartile. The combination of these four coefficients describes a MS.

In spite of the huge differences in complexity of structural and functional connectivity of distant and dissimilar brain regions, compared to activity coupling in our neuronal networks, the preservation of a scale-free connectivity principle and the signs of configuration recurrence suggests that our model is suitable for exploring general properties of connectivity dynamics. Therefore, we next aimed to develop quantitative tools to further study FC dynamics in our hiPSC-derived networks. We modified a method previously designed to evaluate high-level dynamic properties of brain connectivity in SZ patients, through fMRI measurements [14]. To reduce the dimension of the wFC(t) matrix sets obtained from the different networks to a similar and smaller dimension (note that wFC matrices dimension depends on the number of active neurons in a particular network), we ran an independent component analysis (ICA) [27] per network, throughout the temporal dimension of its FC (Figure 3B.II). This computational method allows separation of a complex signal into additive components [27]. Four different and independent FC configurations, defined here as *correlation patterns* (CP), were identified. By expressing each wFC(t) data in terms of these four CP, we obtained four coefficients or “weights”, each one associated to a specific CP. The weights describe how “enriched” is the wFC(t) of each of the different CP (Figure 3B.III). To have a finite number of FC configurations, the weights were discretized into quartiles allowing grouping similar connectivity configurations (Fig 3B. IV). The unique combination of four discretized weights defined a meta-state (MS), borrowing the term coined by Robyn Miller [14].

### FC in SZ is less flexible and slower in rearranging different configurations

By studying neuronal functional relationships in our *in vitro* model, we may address the question of whether FC dynamic alterations observed in SZ patients’ brains by fMRI, can be seen during the development of neural networks, in a completely different spatiotemporal domain. Using the described methodology, we obtained the FC MSs for each network. A set of connectivity variables (listed and defined in Methods) were quantified and differences between- and within-groups were assessed using mixed linear regression modeling. Figure 4 aims to clarify the definition of these variables. Figure 4A illustrates examples of different MSs visited by a network along time. The meaning of the measured FC variables is explained in Figure 4B.

**Figure 4.**
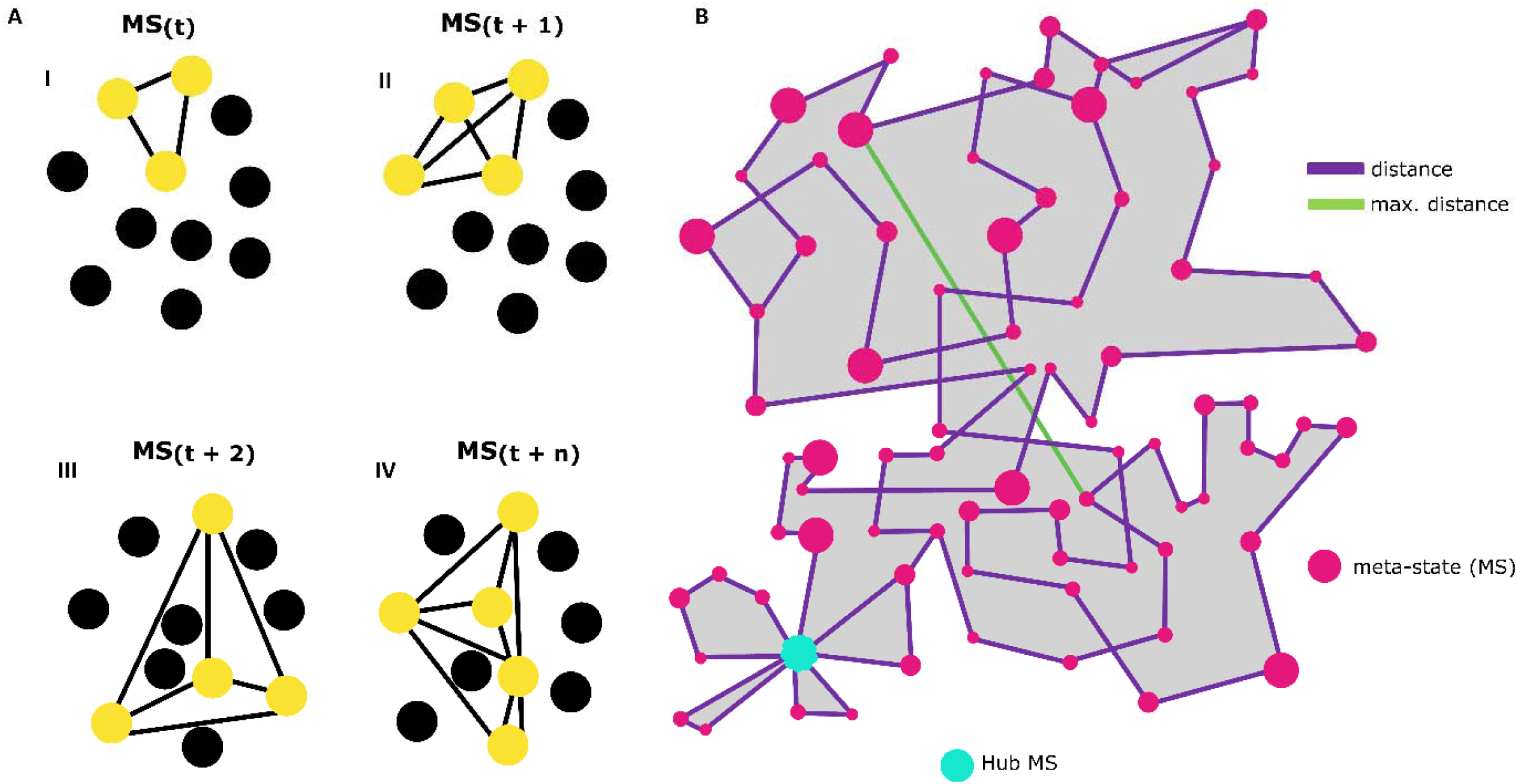
Functional connectivity (FC) flow and FC-related dynamic variables. (A) Schematics representing four possible functional connectivity meta-states (MS), corresponding to specific activity configurations of the neurons conforming the network; each circle represents a neuron. Yellow circles are active, functionally connected (lines) neurons at different times. (B) Functional connectivity flow in a network. Pink circles represent the different visited MSs and circle size denotes the uninterrupted time spent by the network in this configuration. Purple lines represent a distance (Manhattan distance) in the space where the different MSs inhabit. The length of lines connecting MSs illustrates how different are consecutive configurations. Each inflection point corresponds to a transition from an MS to the next one. The green line represents the maximum distance between two successive MSs and illustrates the network’s ability to switch in just one step between two configurations that are as different as possible. The light blue circle indicates an MS that is visited with recurrence, a Hub MS. The grey area corresponds to the dynamic range, a global indicator of the diversity of the accessible MSs. The length of the trajectory of the purple line corresponds to the total traveled distance.

We observed a reduction in the number of different MSs visited by SZ networks (Figure 5A), suggesting a more limited repertoire of accessible configurations, compared to HC networks. We also measured the number of times that networks changed from one MS to another and the mean uninterrupted time spent in a MS. SZ networks change fewer times from one MS to another (Figure 5B) and spend longer at the same MS (Figure 5C), thus they are less dynamic. On the other hand, HC networks can transit to more different (distant) configurations in one step (maximum distance between successive MSs, Figure 5D) than SZ networks. This means that the latter have less potential to fast switching between distant FC configurations, displaying less flexibility. We also calculated the total travelled distance within the MSs-inhabited space, calculated as the sum of the distances between successive MSs through the whole recording time (Figure 5E), finding that it was lower in SZ (Figure 5F). Finally, we evaluated the dynamic range, which is formally a measure of the size of the space containing the different MSs, by calculating the Manhattan distance between the farthest MSs visited along the whole trajectory (Figure 5G). We must bear in mind that the dynamic range calculations are relative to network’s specific CPs. Therefore, this measurement reflects how much the space where the different MSs inhabit is expanded or contracted with respect to the network’s own intrinsic “motion possibility space”. This can be understood as the potential of a network to flow through more diverse connectivity configurations. We observed a reduced FC dynamic range in SZ networks (Figure 5G). This result is consistent with the lower number of different realized MSs and the reduced total traveled distance in SZ networks, as a lower dynamic range implies a decrease in FC motion potential. These findings suggest that resting-state SZ networks exploring available FC configurations less thoroughly and reorganize less rapidly compared to HC. Overall, these results point to a reduction of FC dynamism and flexibility in SZ networks, involving speed and exploration potential.

**Figure 5.**
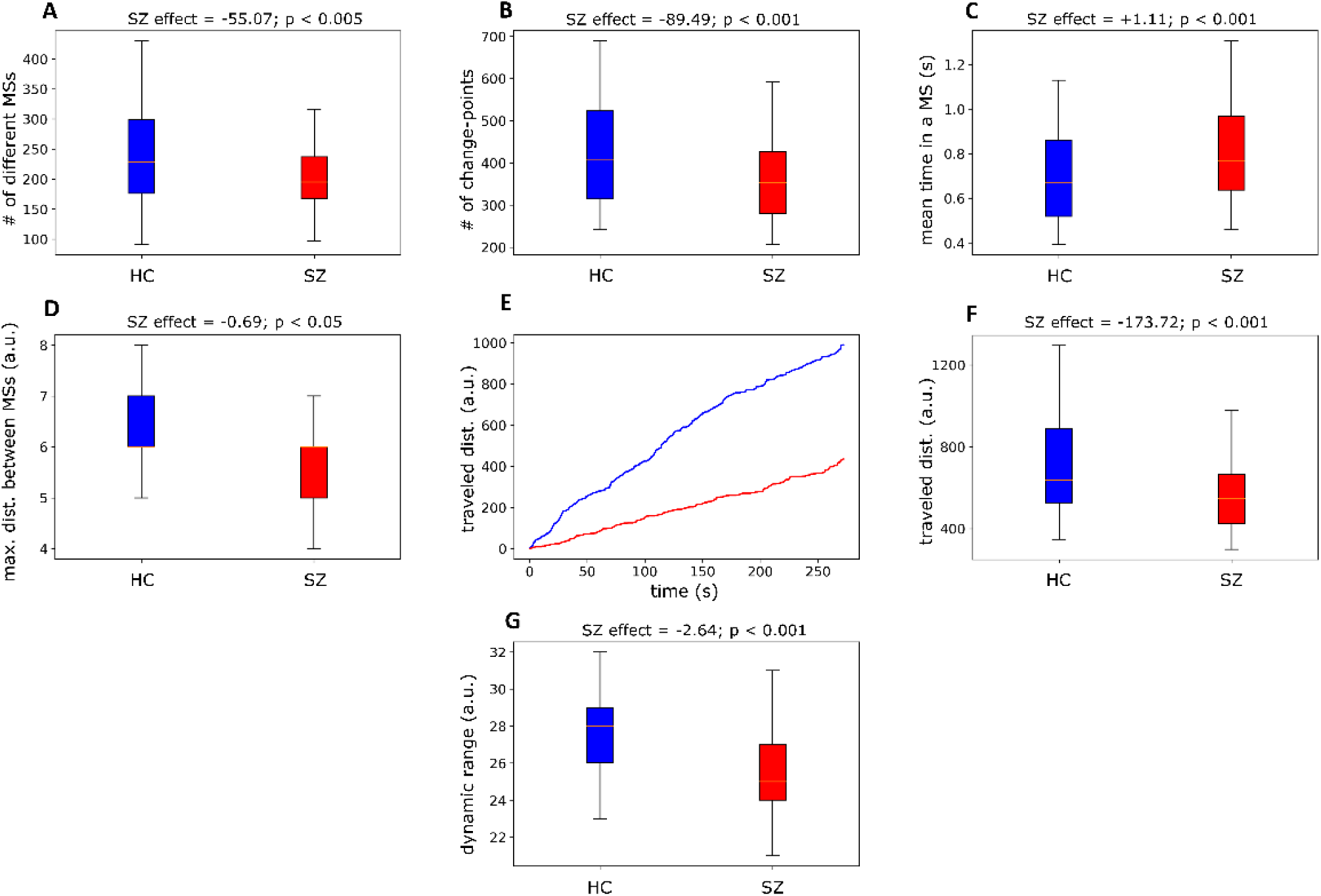
Functional Connectivity in SZ-derived networks is less dynamic than for HC cultures. (A) SZ networks visited a lower number of different meta-states (MSs). (B) SZ networks change fewer times from one MS to another during the recorded time, spending longer uninterrupted time within the same MS (C). (D) The maximum distance between successive MSs is also reduced in SZ neuronal networks. (E) A graphical representation of how FC travels within the space inhabited by the MSs, considering the distance between successive visited MSs. The plot indicates the cumulative traveled distance, measured as the total traveled distance at each time point (x). (F) The overall distance traveled through the MSs space, calculated as the sum of the L1 distances between consecutive MSs, is reduced in SZ networks. (G) The intrinsic dynamic range, measured as the Manhattan distance (L1) between the most distant MSs, is reduced in SZ networks. For each dependent variable (Y-axis), the SZ (*diag*) effect from the regression model, (Materials and Methods), is indicated.

### SZ networks display a reduced number of recurrent FC meta-states (hub meta-states)

As mentioned, neuronal networks went through different FC configurations that reoccur during the acquisition time (Figure 3A, FCD matrix). We computed the number of different MSs that were re-visited by each neuronal network. In whole brain fMRI FC analysis these recurrent MSs have been named as “hub meta-sates” [14]. Our results show that SZ networks display a lower number of hub MSs (Figure 6A) and visit hub MSs less often than HC networks (Figure 6B) but spent longer uninterrupted time in them (Figure 6C).

**Figure 6.**
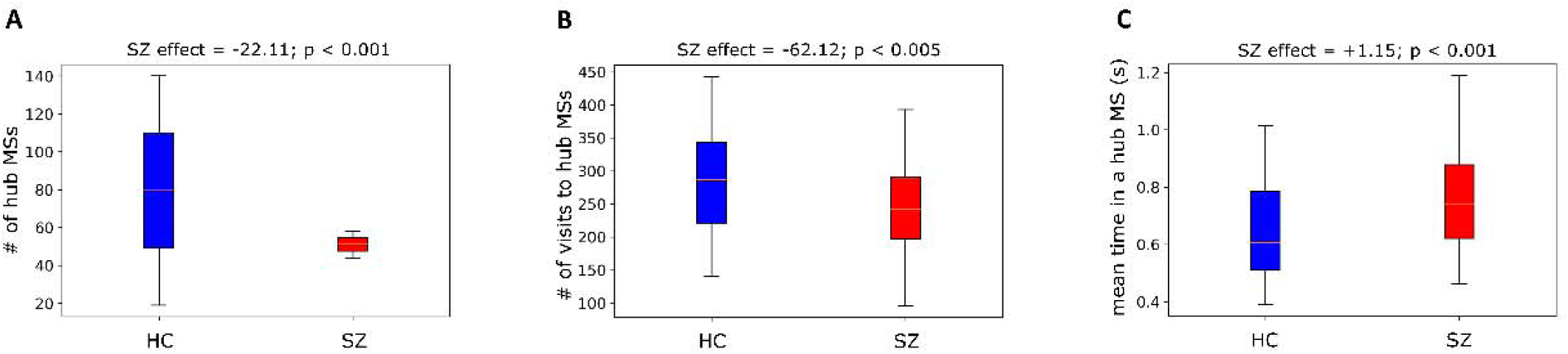
SZ-derived networks visit a lower number of different hub states but remain longer times within them. (A) The number of different visited hub meta-sates (MSs) is lower in SZ networks. (B) SZ networks visit hub MSs less often than HC networks, but (C) spend longer uninterrupted time in the same hub MS. For each dependent variable (Y-axis), the SZ (diagnosis) effect from the regression model, detailed in the Methods and Materials section, is indicated. The p-values of the coefficients meet the requirement of p <0.05.

## DISCUSSION

Cognitive functions require the integration of neural activity through many scales, from neurons and local circuits to large-scale brain networks. This has motivated the recent interest in studying the brain from a perspective of complex systems. Considering the wide evidence for SZ-related FC alterations [7-10, 14], this study aimed to evaluate whether FC differences among SZ patients and HC subjects could be explored during nervous system development *in vitro*, using hiPSCs-derived neuronal networks.

We report that resting-state FC in long-term neuronal cultures is a dynamic but not random phenomenon, presenting recurrent configurations. This resembles what has been widely described at a large scale in human brains by using fMRI, characterized by a much lower spatial and temporal resolution [5]. Hence, addressing the differences and similarities of pattern configuration dynamics across different spatiotemporal domains can contribute to understanding the neurobiological bases of brain integration in both health and disease.

A first interesting point to note is that our results are consistent with the concept of a “default mode” of neural activity, detected in different brain areas in resting conditions and proposed to reflect intrinsic functional properties of neuronal networks, providing an adequate starting point for efficient response to environmental changes [44]. Following this idea is tempting to hypothesize that the altered SZ network activity observed in our study could rely on a distorted network auto-organization, resulting from anomalous neurodevelopmental processes. On the other hand, the scale-free property of both HC and SZ connections, is consistent with the proposal of more general principles ruling brain connectivity organization [45]. However, these possibilities require further evaluation.

Several genes related to neurodevelopment and synaptic function have been implicated in the pathophysiology of SZ. We observed that the expression of genes encoding key glutamatergic postsynaptic proteins presented an altered evolution during early development in SZ networks. The decreased expression of excitatory synaptic components in HC contrasted with the rising trend observed in SZ. On the other hand, the gene encoding the GABAergic synaptic scaffold GEPHYRIN was also strongly downregulated in HC, in contrast to SZ networks, which do not change significantly. Overall, our results suggest an impairment in the excitation/inhibition balance in SZ networks, with a possible tendency to develop hyperexcitability, consistent with observations in SZ brains [8, 32, 46]. The alterations in *CDK5R1, RELN* and *GAD67*, involved in neuronal positioning, network structure and synaptogenesis, are in line with a divergent connectivity evolution during early development in HC and SZ networks [33-37].

Differences in *ATP5* expression patterns could reflect alterations in cellular metabolism during early developmental stages in SZ. Interestingly, recently Hahn et al., 2020 [39] described the intimate relationship between brain metabolic demands and FC during cognitive tasks. It would be of great interest to study at a neuronal level how metabolism changes could modulate FC dynamics.

Synapse number and composition change dynamically during neuronal network shaping across development [47]. Structural and molecular synaptic properties evolve in intercorrelation with network activity, and it is thus not straightforward to interpret which are the key factors leading to an abnormal circuit function at specific developmental stages, particularly in a polygenic disease as SZ. With this in mind, we explored possible fundamental differences in the dynamics of network auto-organization during hiPSCs neurodevelopment, by evaluating functional connectivity variables. We focused on exploring intrinsic features ruling network dynamics during hiPSC neurodevelopment that could be altered in this pathology.

According to our results, SZ networks visited fewer different FC configurations (Figure 5A), presented a smaller number of transitions among MSs (Figure 5 B), and spent longer times in the same MS (Figure 5C). Moreover, the maximum change in a single step that FC configuration can achieve is reduced in SZ networks (Figure 5D), evidencing alterations in FC fast switching and a lower diversity of accessible configurations. The total traveled distance within the space explored by MSs is also reduced (Figure 5F). Finally, the decreased dynamic range indicates that neuronal networks configurations in SZ are more restricted than HC circuits in exploring the limits of their own operative range (Figure 5G). Dynamic range can be considered as a global indicator of the diversity of the accessible MSs. Taken in totality our findings indicate that SZ networks are characterized by a narrower diversity of connectivity configurations, an impaired dynamism, and lower flexibility, compromising the network’s ability for rapid and efficient reorganization. Remarkably, the altered FC in our hiPSC-derived neuronal networks strongly resembles the resting-state dynamics observed in SZ patients’ brains [14]. Miller quantified the same FC dynamical variables, and it is astonishing to conclude that measurements at two such distant spatiotemporal scales are consistent. The differences in neuronal communication reported here could reflect the FC connectivity dynamics of early stages of neurodevelopment in SZ. That is, SZ could be linked to alterations in brain connectivity associated with disease symptoms.

An important observation is the existence of recurrent FC meta-states in hiPSC neuronal networks. SZ networks showed a reduced repertoire of hub meta-states, visit hub MSs less frequently, but remain longer periods of time trapped at the same configuration. It has been proposed that long-term potentiation (LTP; a widely studied model of synaptic plasticity) could constitute the neurophysiological basis for the formation of brain hub states [48]. We hypothesize that the differences we observe may be due to an altered plasticity mechanism of reinforcing certain neuronal connections during neurodevelopment, resulting in fewer different configurations strengthened in the SZ condition. This possibility is supported by the fact that genome wide-association studies [49-51] and a recent single-nucleus RNA sequencing (snRNAseq) analysis in SZ [52], have evidenced alterations in sequence and expression of several genes implicated in specific synaptic pathways related to plasticity [53].

In wakeful rest, as well as during slow-wave sleep, brain activity is characterized by the emergence of spontaneous, temporally compressed episodes (“sharp wave ripples”) of reactivation of groups of neurons that were recruited during recent learning events [54, 55]. These pattern reoccurrences have been associated to the consolidation of synaptic-plasticity-related memories but are also spontaneously generated during resting periods before experience in both animal models and humans [56]. Interestingly, in humans a better memory consolidation correlates with higher repetition frequency during resting intervals (“waking replay”) [57]. Most intriguingly, a very recent study reports alterations in the temporal dynamics of waking replay events in SZ patients, characterized by a reduced replay rate and increased ripple power [58]. Considering the striking similarities between FC dynamics in our *in vitro* model and SZ brains at rest [14], we could speculate a connection among such anomalous replay dynamics and the alterations we observe in the rate and duration of FC reoccurring meta-state (hub MSs). Future research could shed light on the validity of this possibility.

There is an overwhelming amount of rare and *de novo* mutations in SZ, but each of them by itself has only a low effect on disease risk [59, 60]. Our quantitative neuronal secretome analysis did not reveal significant protein expression changes, but interestingly, we do see convergence in a more complex feature as in neurons communicational relationships. The latter leads to easily identify FC dynamic behaviors that differ between conditions. SZ is considered a polygenic disease with multiple etiologies[2], but there is confluence in a variety of symptoms that allows it diagnosis [61]. Thus, it is highly relevant to search for the functional units that may be altered in SZ, rather than focusing on particular molecules, since they can strongly vary from one patient to another.

The brain is a high oxygen- and energy-demanding organ. Thus vascular patterning, blood-brain barrier function, and adequate neuronal energy supply are major factors that ensure its optimal function [62]. Changes in neural activity are sufficient to trigger modifications in vascular network establishment and function [63, 64]. Hence, our reported alterations in neuronal FC dynamics could be directly involved in alterations in the neurovascular unit and the function of the blood-brain barrier, as has been previously suggested for SZ [65-69]. These early neurodevelopmental alterations could converge into a more susceptible organism to harmful or stressful external factors, leveraging it to SZ development [70].

Our results support previous hypotheses suggesting that the brain is a highly complex system that may possess spatiotemporal scale-invariant principles that govern its functioning [5][71]. We propose that altered FC in SZ brains emerges during early neural development, as a consequence of an aberrant neuronal communicational dynamics, jeopardizing the system’s ability for fast and efficient reorganization. Our findings highlight the relevance of identifying alterations in brain functional units, since this may allow to project their repercussions at more complex levels of organization. As per our knowledge, our paper represents a first contribution to this challenging effort.

## Data Availability

All the data are available upon request. All proteome membranes are displayed in annexed figures.
Raw neural connectivity data along with code that enables analysis and measurement of FC dy-namics available at https://doi.org/10.5281/zenodo.5130447

https://doi.org/10.5281/zenodo.5130447

## AUTHOR CONTRIBUTIONS

S.P. differentiated cell cultures in collaboration with B.C., completed imaging experiments, wrote code, analyzed imaging data, implemented the mathematical modelling of *in vitro* FC dynamics and wrote the first draft of the manuscript. K.B. built the imaging set-up, designed and participated in the imaging experiments, and conducted the electrophysiological recordings. R.M., oriented S.P. to adapt her published mathematical methods for whole-brain dynamics to a cellular model and supported with data analysis. B.C. and D.G. designed, performed and analyzed qPCR and proteomic experiments. All authors actively participated in the interpretation of results and edited and improved the manuscript. M.S. contributed to research design, imaging setup implementation, guidance of mathematical analysis and manuscript redaction. Conceptualization, formal analysis writing-review and editing, and general supervision was conducted by V.P.

## ACKNOWLEDGEMENTS

We are grateful to Dr. Stevens Rehen (D’Or Institute for Research and Education (IDOR), Brazil for donating cell lines and valuable reagents. This work was supported by: FONDECYT # 119083 (VP), CONICYT Fellowships for PhD studies # 21181102 (SP), CONICYT/ANID Fellowships for PhD # 21150781 (BC).

## AVAILABILITY OF SUPPORTING DATA

All the data are available upon request. All proteome membranes are displayed in annexed figures. Raw neural connectivity data along with code that enables analysis and measurement of FC dynamics available at https://doi.org/10.5281/zenodo.5130447

