## Supplementary tables and figures for "Altered resting-state functional connectivity in hiPSC-derived neuronal networks from schizophrenia patients"

**SUPPLEMENTAL INFORMATION**

**Table S1. hiPSC cell lines used in this study.**

| **Code** | **Cell line** | **Diagnosis** | **Sex** | **Age (range)** | **Cell Source** | **Reprogramming Technique** |
| --- | --- | --- | --- | --- | --- | --- |
| HC#1 | GM23279A | Control | F | 36-40 | Fibroblast | Cytotune 1.0 kit (ThermoFisher) |
| HC#2 | CF2 | Control | M | 31-35 | Fibroblast | Cytotune 1.0 kit (ThermoFisher) |
| HC#3 | ADHD2 | Control | M | 31-35 | Urine Endothelial | (OCT4, SOX2, KLF4, MYC) |
| SZ#1 | GM23760B | Schizophrenia | M | 26-30 | Fibroblast | Cytotune 2.0 kit (ThermoFisher) |
| SZ#2 | GM23761B | Schizophrenia | F | 26-30 | Fibroblast | (OCT4, SOX2, KLF4, MYC, LIN28) |
| SZ#3 | EZQ3 | Schizophrenia | M | 41-45 | Fibroblast | Cytotune 2.0 kit (ThermoFisher) |
| SZ#4 | EZQ4 | Schizophrenia | M | 41-45 | Fibroblast | (OCT4, SOX2, KLF4, MYC, LIN28) |

Information about diagnosis, sex and age of the patients [1,2,3,4]

**Table S2. Specifications of the mixed effect regression models and their output (estimated SZ effects and associated p-values)**

| **Dependent variable** | **estimated SZ effect** | **p-value** | **Optimizer** | **Included predictors** |
| --- | --- | --- | --- | --- |
| # of active neurons | -6.72 | 5.27 x 10^-1^ | BFGS | diagnosis, a random intercept per cell line. |
| # of different MSs | -55.07 | 4.59 x 10^-3^ | CG | diagnosis, # of neurons, # of neurons^2^, a random intercept per cell line. |
| dynamic range (a.u) | -2.64 | 2.57 x 10^-4^ | BFGS | diagnosis, # of neurons, # of neurons^2^, a random intercept per cell line. |
| # of change-points | -89.49 | 9.53 x 10^-11^ | BFGS | diagnosis, # of neurons, # of neurons^2^, a random intercept per cell line. |
| mean time in a MS (s) | 0.17 | 7.48 x 10^-7^ | BFGS | diagnosis, # of neurons, # of neurons^2^, a random intercept per cell line. |
| max. dist. between MSs (a.u) | -0.69 | 2.68 x 10^-2^ | BFGS | diagnosis, a random intercept per cell line. |
| traveled dist. (a.u) | -173.72 | 1.12 x 10^-9^ | BFGS | diagnosis, # of neurons, # of neurons^2^, a random intercept per cell line. |
| # of hub MSs | -22.11 | 2.40 x 10^-5^ | BFGS | diagnosis, # of neurons, # of neurons^2^, a random intercept per cell line. |
| # of visits to hub MSs | -62.12 | 2.73 x 10^-3^ | BFGS | diagnosis, # of neurons, # of neurons^2^, a random intercept per cell line. |
| mean time ina hub Ms (s) | 0.17 | 2.31 x 10^-5^ | CG | diagnosis, # of neurons, # of neurons^2^, a random intercept per cell line. |

SZ effect and p-values for the different variables describing functional connectivity (FC) dynamics, obtained by mixed linear regression modeling (see Methods and Materials). The SZ effect is the coefficient for the diagnosis variable (coded as “1” for SZ and “0” for HC networks). The optimizer and predictors used in each regression model are also shown.

**Table S3. Changes in mRNA expression levels along neuronal differentiation.**

| **Gene** | **mean log fold-change (90/30 D) in HC networks** | **adjusted p-value** | **change (HC)** | **mean log fold-change (90/30 D) in SZ networks** | **adjusted p-value** | **change (SZ)** |
| --- | --- | --- | --- | --- | --- | --- |
| *HOMER1* | -2.07 | 1.38 x10^-5^ | Decrease | 2.30 | 1.14 x10^-3^ | Increase |
| *GRIN1* | -7.36 | 7.07 x10^-7^ | Decrease | 2.28 | 1.19 x10^-1^ | No change |
| *GPHN* | -6.61 | 1.08 x10^-127^ | Decrease | -3.37 | 2.17 x10^-4^ | Decrease |
| *SYP* | 1.26 | 4.26 x10^-1^ | No change | — | — | — |
| *CDK5* | -8.33 | 2.81x10^-77^ | Decrease | -2.38 | 1.59 x10^-2^ | Decrease |
| *RELN* | 4.25 | 5.6 x10^-2^ | No change | -7.55 | 1.62 x10^-2^ | Decrease |
| *GAD67* | -1.86 | 6.13 x10^-2^ | No change | 3.79 | 1.7 x10^-2^ | Increase |
| *GLUT1* | -3.08 | 5.57 x10^-2^ | No change | 2.63 | 1.46 x10^-1^ | No change |
| *SEMA3A* | 1.81 | 1.01 x10^-1^ | No change | 1.10 | 4.32 x10^-1^ | No change |
| *ATP5* | -1.51 | 4.57 x10^-54^ | Decrease | 0.14 | 6.8 x10^-1^ | No change |

Change in mRNA expression levels of different neurodevelopment-related genes, within a period ranging from 30 to 90 days in culture, for HC and SZ conditions. We estimated the mean log2 fold-change in mRNA expression with an unconditional linear model with a random intercept for cell line identity to account for variability within and between the different lines (three SZ (#1, 2, and 3) and two HC (#2 and 3), all in duplicates). The p-value associated with the mean is obtained from the z-score of the estimated intercept in the linear regression model. If the mean log fold-change 90/30 is significantly larger than zero, we considered an increase in gene expression and vice-versa. If p-value > 0.5, we cannot infer any change during the explored period.

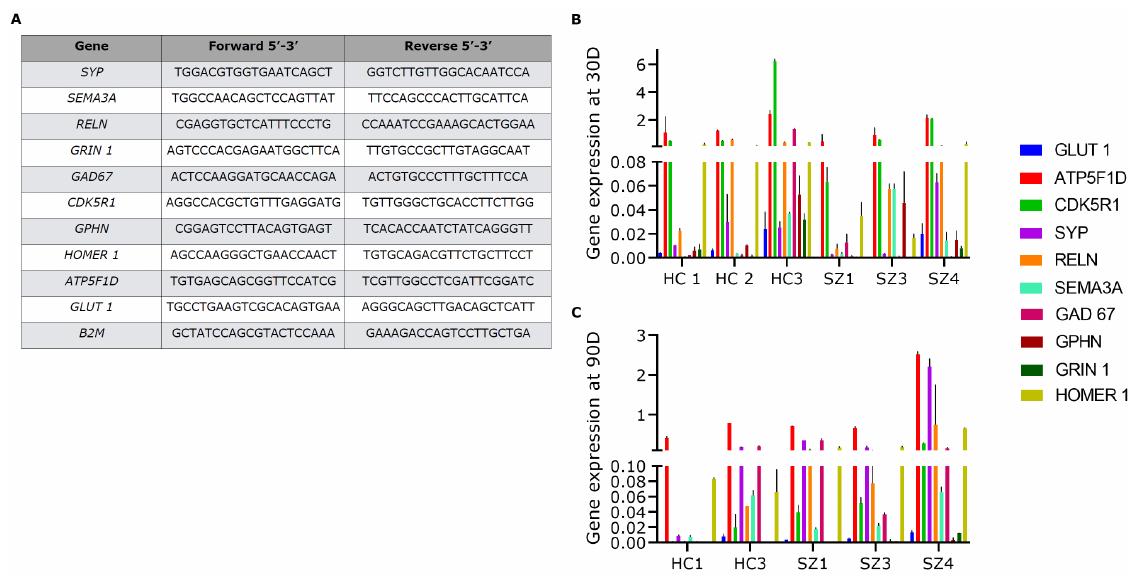

**Figure S1. qPCR analyses of neuronal cultures at 30 and 70-91 days of differentiation.**

(A) Primers used for qPCR amplification. mRNA expression levels of different genes related to nervous system development at 30 (B) and 70-91(C) days of differentiation. *B2M* was used as housekeeping gene. Data are shown as mean ± SD.

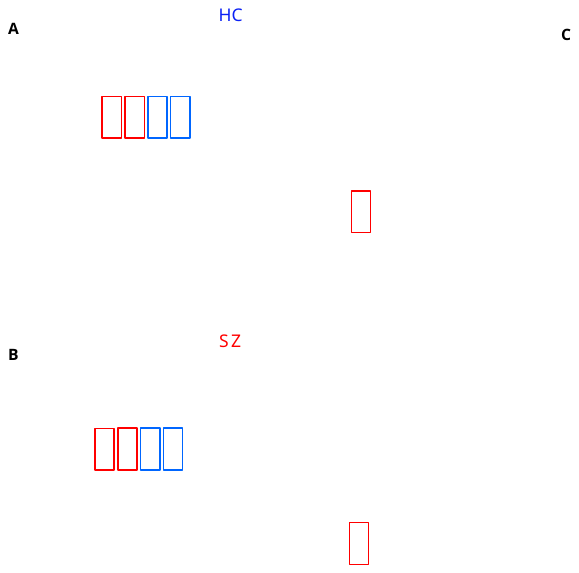

**Figure S2. Neurogenic secretome profiling of hiPSC-derived cultures.** (A-B) Representative pictures of neurogenic secretome profile from HC and SZ conditioned media. Red boxes indicate positive internal control spots; blue boxes show negative internal control spots. (C) Heatmap depicting the levels of neurogenic proteins present in four SZ (#1-4) and three HC (#1-3) cell lines at 75 days of differentiation. Data are shown as average level relative to internal control

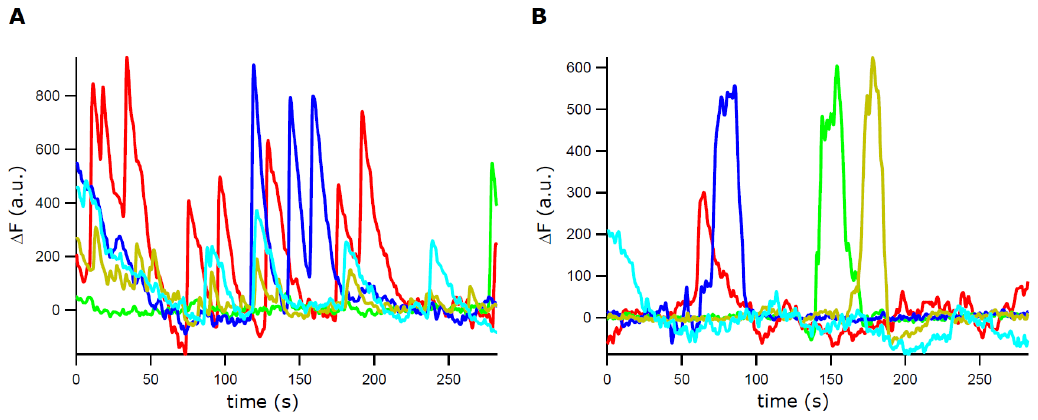

**Figure S3. Ca^2+^ signals depend on activation of voltage-dependent Na^+^ channels.** (A) Ca^2+^ signals of 5 randomly selected neurons from a given network. (B) Reduction in neuronal activity after adding TTX (0.2 µM) to the bath of the network showed in (A). This indicates that the registered activity in the network is due to the activation of voltage-dependent sodium channels in these neurons.

**Figure S4. FC variables correlate to the number of active neurons in a network**. (A) There is no difference in the number of active neurons per network among HC and SZ conditions. (B) Relationship between the different dependent variables and the number of active neurons (B.I-B.IX). In most cases, dependent variables correlated to the number of active neurons in both HC and SZ conditions.

**Annexed Figure.** Proteome membranes of all original data

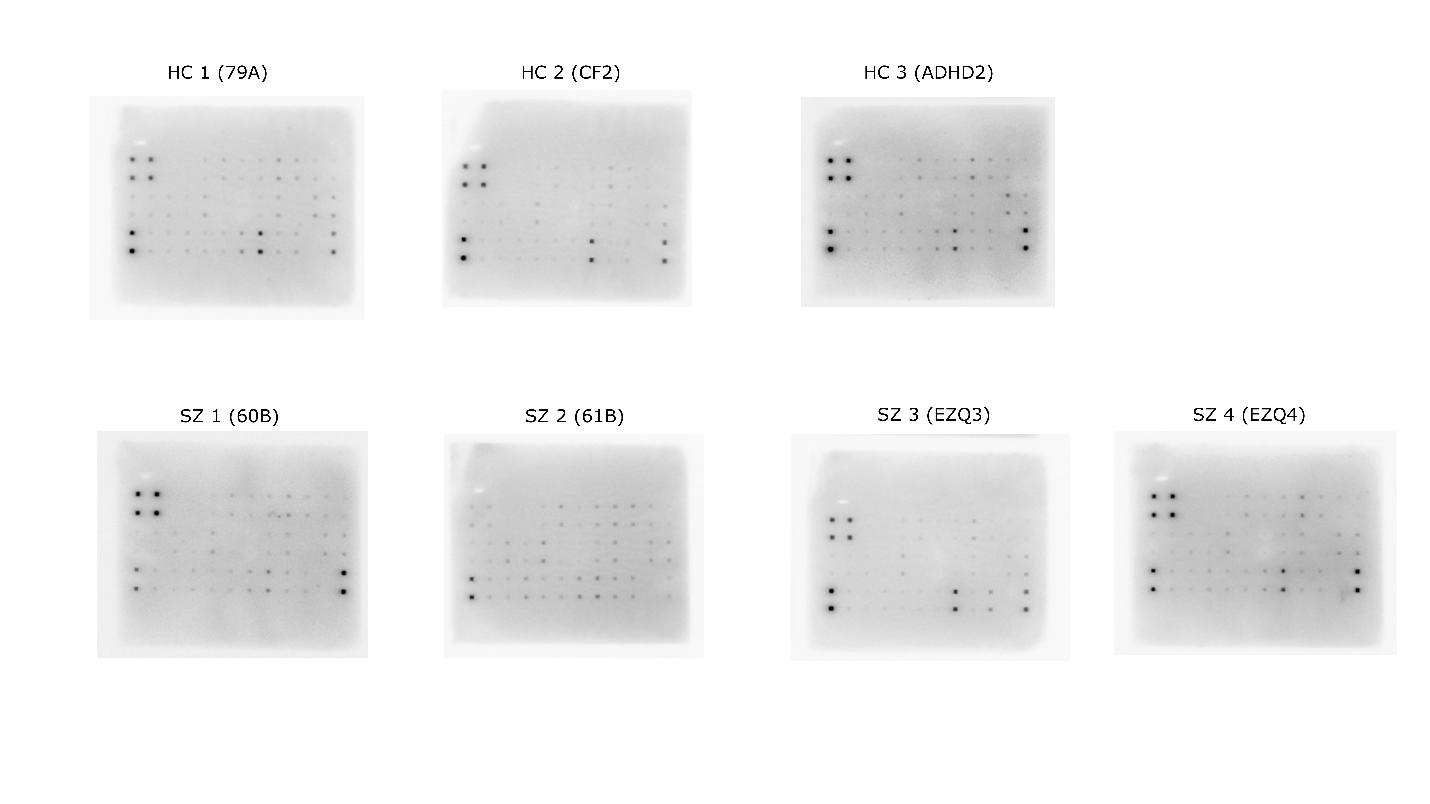
